# Nationwide rollout reveals efficacy of epidemic control through digital contact tracing

**DOI:** 10.1101/2021.02.27.21252577

**Authors:** Ahmed Elmokashfi, Joakim Sundnes, Amund Kvalbein, Valeriya Naumova, Sven-Arne Reinemo, Per Magne Florvaag, Håkon Kvale Stensland, Olav Lysne

## Abstract

Fueled by epidemiological studies of SARS-CoV-2, contact tracing by mobile phones has been put to use in many countries. A year into the pandemic, we lack conclusive evidence on its effectiveness. Here, we used a unique real world contact data set, collected during the rollout of the first Norwegian contact tracing app in the Spring of 2020, to address this gap. Our dataset involves millions of contacts between 12.5% of the adult population, and enabled us to measure the real-world app performance. The technological tracing efficacy was measured at 80%, and we estimated that at least 11.0% of the discovered close contacts could not be identified by manual contact tracing. The overall effectiveness of digital tracing depends strongly on app uptake, but significant impact can be achieved for moderate uptake numbers. Used as a supplement to manual tracing and other measures, digital tracing can be instrumental in controlling the pandemic. Our findings can thus help informing public health policies in the coming months.

---

When the SARS-CoV-2 virus started spreading globally, many initiatives for the development of digital contact tracing based on mobile phones were launched^1,2^. The efforts were motivated by a study by Ferretti et al., which suggested that an effective widely adopted digital contact tracing system may be enough to keep the reproduction number below 1^3^. Almost a year into the pandemic, we have still not seen conclusive evidence that digital contact tracing can play a significant role in containing the pandemic. As a result, several studies have questioned the efficacy and need for digital contact tracing, especially when considering its encroachment on privacy^4–10^. Measuring the effect of digital contact tracing has been notoriously hard, as there is no contacts dataset available from a full scale production system. Further, most of the deployed systems are based on the Exposure Notification System (ENS)^11^, which is designed to greatly limit visibility into contact events in order to preserve privacy. Hence, current assessments of ENS-based apps have resorted to using a combination of incomplete data that ENS provides and population surveys^12–14^. Here, we used a unique real world contact data set, that was collected and anonymized during the rollout of the first Norwegian contact tracing app (Smittestopp) in the Spring of 2020^15^, to tackle these limitations. Our dataset involves millions of contacts and enabled us to measure the real-world technological tracing efficacy of the app, apply a machine learning classifier to estimate the number of contacts not identifiable by manual contact tracing (see Figure 1a) and to parameterize a model that relates tracing efficacy to the app uptake in the population. Finally, we used our efficacy estimates as an input to an established model of pandemic spreading, to assess the potential impact as a control measure.

**Figure 1:**
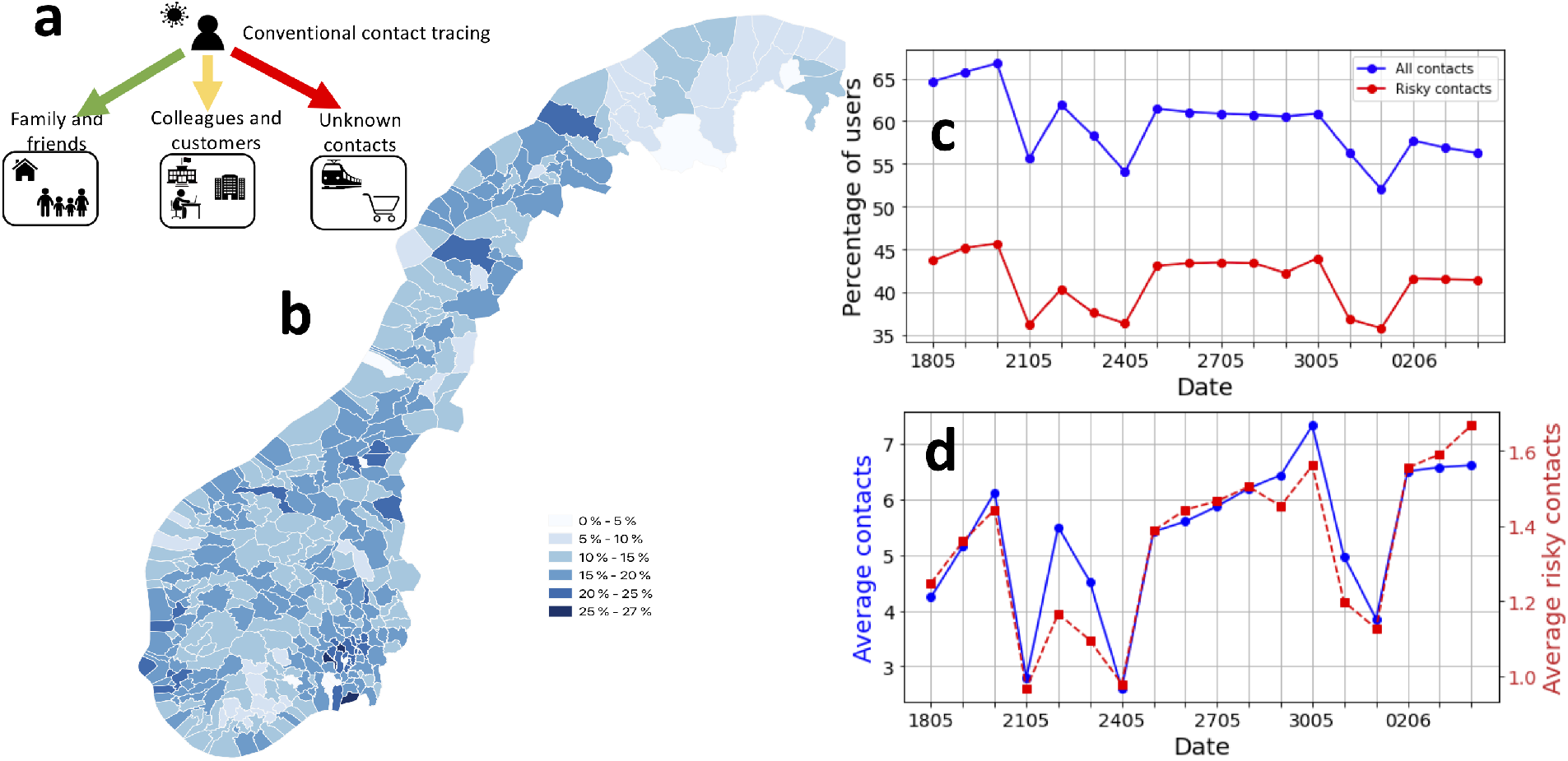
**a)**: Typical settings of social contacts, the colors of the arrows capture whether manual contact tracing can succeed in identifying the contacts in the respective setting (green means complete identification, yellow partial identification, while red means zero or minor identification), **b)**: The percentage of population over 16 year old that were using Smittestopp in each municipality, **c)**: the percentage of active users per day as well as the percentage of active users that were involved in risky close contacts (within 2 meters) that lasted 15 minutes or more, **d)**: The average number of contacts per day over time, all contacts (blue) and risky close contacts (red).

We measured a high success rate in accurately detecting nearby devices (80%). Further, we estimated that a non-trivial percentage of the traced close contacts were not visible to manual contact tracing (at least 11%). We also found that the overall effectiveness of digital tracing is strongly dependent on app uptake. While an overall tracing efficacy comparable to manual contact tracing requires app uptake in the range from 80% to 90%, we found that significant impact can be achieved for much lower uptake numbers. For example, an uptake of 40% would be enough, assuming a fast and effective case isolation, for controlling a pandemic with reproduction number of 1.5. Our results add to the emerging evidence that apps are a valuable public health tool^12–14^.

## Digital contact tracing at scale

Smittestopp was rolled out in the Spring of 2020 and was quickly installed by 28% of the adult population (see Figure 1b). The app was eventually suspended in June, because of a combination of low infection rates and privacy concerns^16^. An ENS-based app was launched in December 2020^17^. Smittestopp used Bluetooth low energy (BLE) to discover phones in a range of 10 meters. Upon a discovery event, the app measured the power of the received BLE signal, which was used to approximate the distance to the discovered device. The devices would upload their measurements to a central server, which fused the received data for identifying contacts. This centralization ensured a symmetric contact identification, since Smittestopp was asymmetric by design, that is a detection event in one direction does not imply the opposite is true.

To track the effectiveness of Smittestopp, we used anonymized daily aggregates of BLE discovery events, contacts, spanning 18 days (see the Supplementary Information). We recorded over 26 millions contacts between 545354 phones (i.e., 12.5% of the adult population in Norway). The percentage of daily active users fluctuated between 50% and 70%. Two thirds of the daily active users were involved in a single risky contact (see Figure 1c), which corresponds to being within 2 meters from another person for 15 minutes or longer ^18^. We also found that 80% of the active users had contacts on at least five different days. When considering the entire data set we found that 89.6% of users had at least a single risky contact that lasted 15 minutes or longer. The observed retainability of the app and the pervasiveness of close contacts suggest a reasonable case coverage, that is the fraction of positive cases using the app. Assuming a homogeneous uniform mixing between app users and the rest of the population, we expect a case coverage close to the app adoption level.

To investigate whether Smittestopp captured movement patterns in the society, we examined the number of contacts and risky contacts over time (see Figure 1d). Both numbers exhibited a slightly increasing trend, which is consistent with the fact that society was slowly opening up during this period. The average number of contacts dropped in weekends and national holidays. The trend of risky contacts followed closely that of all contacts. The development in the average contacts matches well known properties of human contacts ^19^.

## Estimating the technological efficacy

We used the collected contact events to estimate the tracing efficacy of Smittestopp, the probability that a physical proximity event between two phones is detected by the app, and how it is impacted by app uptake. The key assumption to computing the efficacy of Smittestopp is that all phones detect each other independently. The global mobile phone market is dominated by two operating systems; iOS and Android, which we have found to differ significantly in their ability to detect contacts. We let *T* be the set of all unique unordered pairs of phone architectures, and instantiate our model by letting *T* = {[*i, i*], [*a, a*], [*i, a*]} where *i* means iOS, and *a* means Android. Using the collected contact events, we estimated the following probabilities: *p*_*ii*_ = 0.54 (iPhone detects iPhone), *p*_*ai*_ = 0.53 (Android detects iPhone), *p*_*ia*_ = 0.53 (iOS detects Android), and *p*_*aa*_ = 0.74 (Android detects Android). These probabilities remained stable throughout the measurement period (see Figure 2a). The details of the underlying assumptions and calculations are provided in the Methods section.

**Figure 2:**
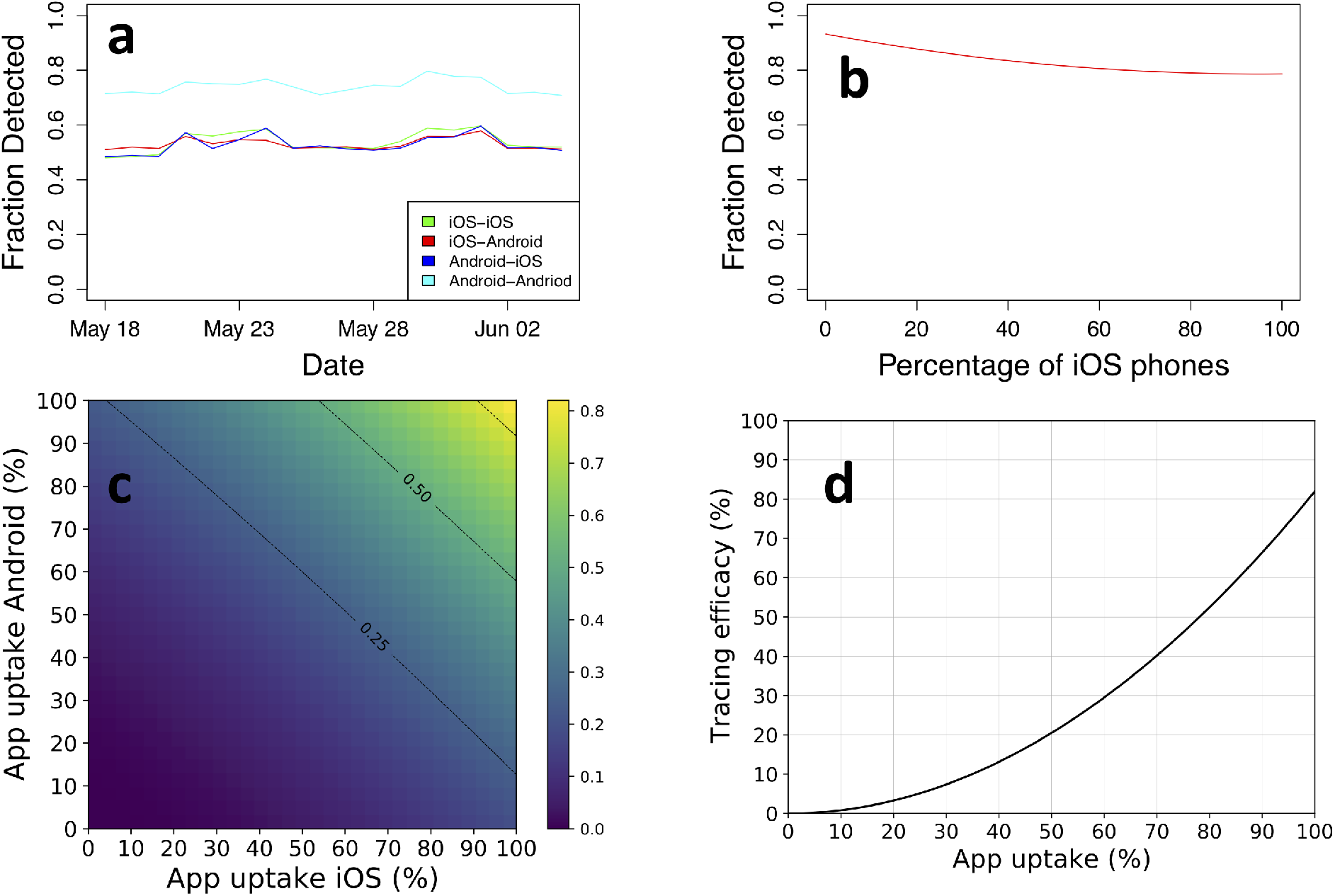
**a)**: The probabilities of detection between different pairs of architectures as it developed over a period of 18 days, **b)**: The detection of contacts varies between 93% with only Android phones in the population to 79% with only iOS phones in the population. The detection rate is 80% when we have an equal split, **c)**: The efficacy of tracing as a function of app uptake in the two user groups. The lines mark different iOS market shares, **d)**: Tracing efficacy as a function of app uptake, assuming the same uptake in the two groups as well as an equal market share 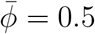.

Assuming a full app uptake in the population, the tracing efficacy of a centralized architecture, *E*, can be formulated as

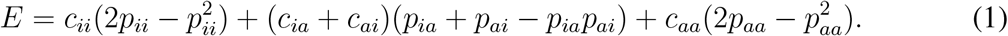

Here, *c*_*x*_ denotes the probability that a physical contact between two phones is of type *x* ∈ {*aa, ii, ai, ia*}. The four values for *c*_*x*_ can be calculated directly from the fraction of the different operating systems of the phones using the app. If we define *M*_*i*_ = *ϕ* as the proportion of apps running on iOS phones, and *M*_*a*_ = 1 − *ϕ* as the proportion running on Android, we have

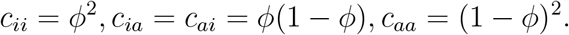

These calculations show that the efficacy of the system depends on the distribution of iOS and Android phones in the population. The theoretical efficacy of the system in terms of detection of contacts varies between 93% with only Android phones in the population to 79% with only iOS phones in the population, as illustrated in Figure 2b.

In reality the uptake of contact tracing apps is well below 100%, and for Smittestopp we also observed that the uptake differed significantly between iOS and Android users. To give a realistic estimate of the app tracing efficacy, we needed to modify (1) to incorporate these factors. We define *α*_*i*_, *α*_*a*_ as the app uptake among iOS and Android users, respectively. Then (1) still holds, but with modified values for the probabilities *c*_*x*_, *x* ∈ {*aa, ii, ai, ia*}:

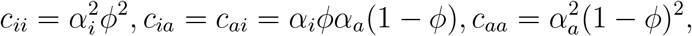

Figure 2c shows the tracing efficacy as a function of app uptake among the iOS and Android users, calculated using these modified *c*_*x*_ values with (1) and the detection probabilities *p*_*x*_, while Figure 2d shows the tracing efficacy assuming an equal uptake by the two groups. The overall effectiveness of digital tracing is strongly dependent on app uptake, and follows the expected quadratic curve. The provided expressions for technological efficacy can be applied to systems other than Smittestopp given that the terms in Eq. 1 can be estimated. Our formulation of the technological efficacy gives an estimate of false negatives produced by the system, but it does not capture false positives. Given the modest secondary attack rate of the virus^20,21^, we expect technological false positives to have a minimal impact on the number of wrongly isolated cases (see the Supplementary Information).

One weakness of our dataset is that it was collected from an app that was developed before the ENS, the current de-facto standard for digital contact tracing, became available. It must be noted that although ENS is expected to perform better than Smittestopp, this has not been possible to verify in any deployed system. The technical reasons for this are presented in Supplementary Information. Limited experiments in controlled environments do, however, support the assumption that ENS will have an efficacy comparably to or better than we have observed in the deployment of Smittestopp^22,23^, and thereby support our conclusions on the potential of digital contact tracing. The available followup data on deployed ENS-based apps is limited. We used publicly available statistics about the German and Swiss official apps to gauge their efficacy^24,25^. These two apps were rolled out in June 2020. Inline with our results from Smittestopp, the app uptake seems to be a good proxy for gauging the case coverage (see the Supplementary Information).

## Detecting unknown close contacts

To check whether Smittestopp was successful in detecting untraceable close contacts, we built a machine learning classifier to separate unknown (random) contacts from known close contacts (see the Methods). The model learned association patterns from the contacts graph and achieved an accuracy of 89% when classifying risky close contacts. Overall, at least 11% of the risky close contacts were random.

The fraction of daily random contacts varied slightly over time, but remained around 6% (see Figure 3a). It dropped during holidays, notably the long Ascension day weekend in the end of May, and it peaked in the days leading to the holidays. We performed the same analysis as we varied the threshold for considering a contact as risky. The fraction of risky random contacts increased to 33.3% when abolishing the duration threshold. We, however, note that our estimates of random contacts are conservative (see the Methods and Supplementary Information).

**Figure 3:**
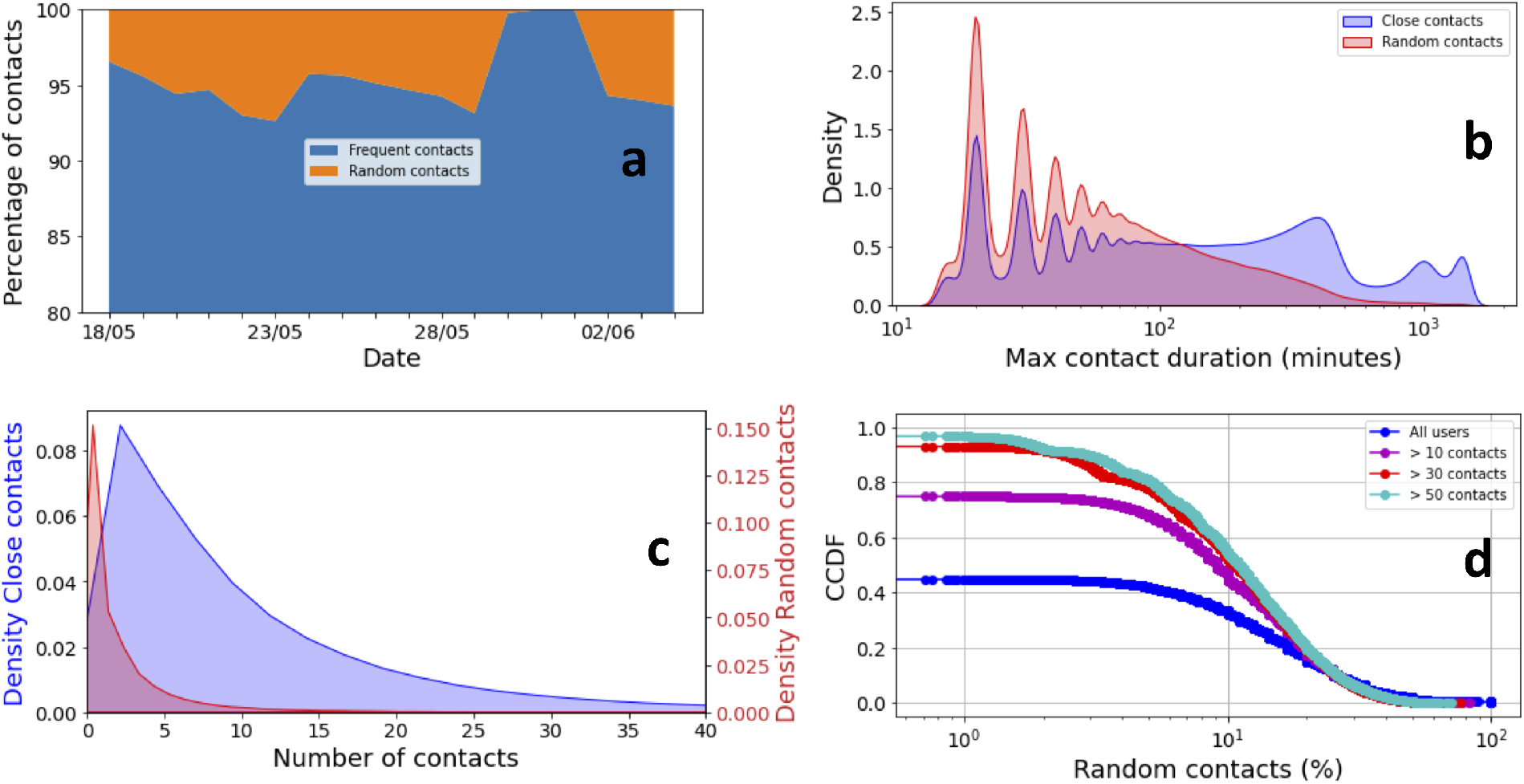
**a)**: The contacts split over time. The fraction of risky random contacts is stable over time and decreases in weekends and holidays, **b)**: The density of encounter duration. Frequent contacts tend to be markedly longer, **c)**: The density of the number of contacts per user, which are split based on the inferred contact type, **d)**: The complementary cumulative distribution of the percentage of random contacts per user for all users, users with 10 contacts or more (49% of all users), users with 30 contacts or more (12% of all users) and users with 50 contacts or more (4% of all users).

Random contacts were shorter, mostly lasting between 20 and 40 minutes (see Figure 3b). This is, however, a duration long enough to spread infection. Close contacts were longer and can last a full day (i.e. household contacts) and several hours with a peak around 7 hours (i.e. work contacts). The number of random contacts per user, in the entire study period, was far less than close contacts. Over half of the users did not have random contacts (i.e. only known contacts), but over 30% of the users had 10% or more random contacts (see the lower Figures 3c and Figures 3d). The lack of random contacts, for over 50% of the users, can be related to the imposed lockdown and adherence to social distancing as well as our conservative estimates. Considering only users with a relatively high number of contacts, the percentage of users with 10% random contacts increased to between 50% and 60%. Overall, the top 20% of users, in terms of contacts, had 20% or more random contacts. This suggest that digital contact tracing can potentially help containing super-spreaders.

The detected fraction of random contacts suggests that the app can significantly supplement manual contact tracing. If we assume, for instance, 60% app uptake in the population, we observe from Figure 2d that the efficacy of the app tracing is approximately 30%. This app will improve the overall tracing accuracy by 7.5% to 10.5% in a society where the fraction of random contacts is between 25% and 35%, respectively. Here, we assume that manual contact tracing identifies all non-random contacts. A supplement of this magnitude can mean the difference between a controlled pandemic and an exponential growth of cases^3^.

## Effect on spread of SARS-CoV-2

After investigating the technological efficacy and its viability in detecting contacts, we turned to assess its potential impact on the pandemic spread. The tracing efficacy as a function of app uptake was computed from (1), as illustrated in Figure 2c, and then these numbers were input to the model of Ferretti et al^3^, which describes the effect of contact tracing on pandemic spread. Figures 4a and 4b show the estimated growth rate *r* (in days^−1^), as a function of app uptake in the two user groups. In Figure 4a, we chose the initial reproduction number as *R*0 = 2.7, which is in line with reported numbers from the early phase of epidemic spread in various countries^26,27^. Figure 4b shows the growth rate for *R*0 = 1.5, chosen to represent a more slowly growing epidemic resulting from control measures such as social distancing. In both plots, the efficacy of isolating symptomatic cases was set to 70%, we assumed a four hour delay in both case isolation and quarantining of contacts, and the proportion of environmentally transmitted (i.e., non-traceable) infections was set to 10% inline with our estimates. Results for other parameter choices are included in the supplementary material. The black lines show *r* = 0, i.e., the threshold between increasing and declining numbers of infected cases. Figure 4c shows the same results as curve plots, assuming identical app uptake in the two user groups.

**Figure 4:**
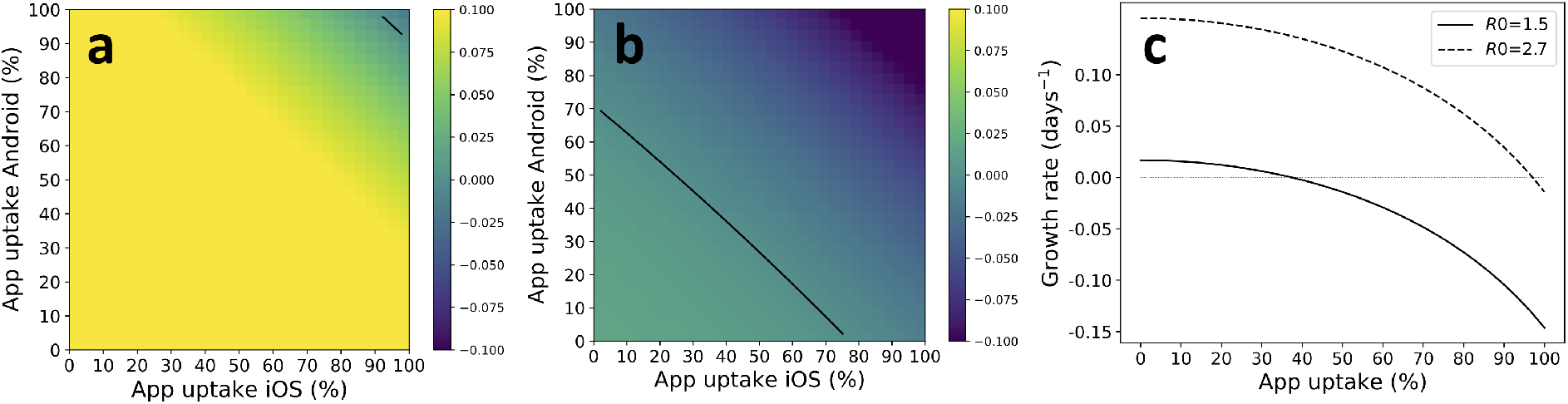
The plots show the estimated growth rate *r* as a function of app uptake among Android and iOS users. We have assumed 90% efficacy of case isolation and a 4 hour delay of both case isolation and contact quarantining. **a)** shows the situation for *R*0 = 2.7 and **b)** shows *R*0 = 1.5. **c)** shows the same data, but assuming identical app uptake among iOS and Android users.

Figure 4a indicates that controlling the pandemic using app-based contact tracing alone is probably unrealistic, since 95% of the population would need to install the app to control a pandemic with an initial reproduction number *R*0 = 2.7. However, the situation is completely different for the case of *R*0 = 1.5, which may be more representative for a situation with other controlling measures in place. In this case, the required uptake was about 40% to control the epidemic and achieve a decline in the number of cases. Unfortunately, the majority of countries struggle with pushing app uptake beyond 20%-25%. However, a few countries including the UK and Denmark, provide a cause for optimism, by reaching a 30% or more uptake rate. For example, 49% of the eligible population with compatible phones have installed the NHS app in England and Wales^14^.

## Discussion

Our analysis of the data from the first rollout of Smittestopp reveals some central findings. The first is that it is possible to reach a significant efficacy of digital contact tracing on mobile phones. With an equal split between Android and iOS-phones in the population, we measured an efficacy above 80%. Although many discussions on the topic have taken this for granted, it should be noted that this was far from obvious. The phone-models used in a given society varies enormously, and none of the models were designed with contact tracing in mind. Documentation of an efficacy of above 80% in a rolled out solution is therefore a decisive finding. We expect ENS-based apps to achieve a comparable or better accuracy, given their superiority to Smittestopp. The second finding is that when used in a real population, a digital contact tracing system does detect a non-trivial number of close contacts that is out of reach for manual contact tracing. Our machine learning model concluded that at least 11% of the contacts with high risk of infection spread were random, and would likely not have been identified with manual contact tracing. The third finding is that measurements from a full scale rollout combined with epidemiological models show significant potential contributions from digital contact tracing to stopping the pandemic. Although there is significant room for improvement of technical accuracy, it appears that the app uptake rate in the population is the only real impediment for realizing the potential of digital contact tracing. Yet, tangible benefits are possible at modest uptake rates. An uptake rate of 40%, for example, can help reducing the reproduction number in the current phase of the pandemic, where the reproduction number is between 1 and 1.5 in most countries^28,29^.

With Covid-19 expected to be endemic^30^, new virus variants^31^ and a promising but lengthy vaccination campaign^32^, our findings suggest that digital contact tracing can greatly boost efforts to control the pandemic in the coming months. Health authorities should have an immediate focus on increasing the uptake of contact tracing apps. This should be supplemented with efforts to establish digital contact tracing as an essential tool for public health.

## Methods

### Modelling the technological efficacy

A central assumption in the model is that the phones detect each other completely independently. This means that whenever phone *A* detects phone *B*, this detection event does not change the behavior of phone *B* in a way that will affect its probability of detecting phone *A*. There is nothing in the implementation of Smittestopp that should imply that this assumption does not hold. The code was written such that the act of detecting another phone, and the act of being detected by another phone are not dependent on each other.

Assume two types of phones, *x* and *y*. When two such phones are in proximity of each other, let *p*_*xy*_ be the probability that *x* detects *y*, and *p*_*yx*_ be the probability that *y* detects *x*. Let *C*_*xy*_ be the number of factual proximity events, i.e., contacts, over a given period, between two app users carrying phone of type *x* and *y*, respectively. Furthermore, let *D*_*xy*_ and *D*_*yx*_ be the number of these events that are detected by the phone of type *x* and *y*, respectively, and let *D*_*xy*+*yx*_ be the number of proximity events detected by both phones.

Then the following equations hold:

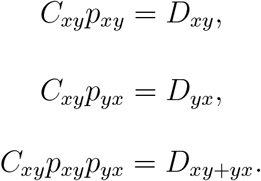

Solving these equations for *p*_*xy*_ and *p*_*yx*_ gives

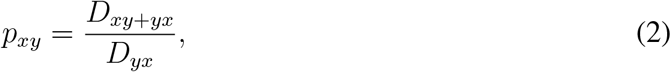

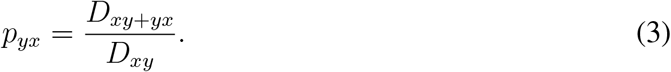

Note that these equations are valid regardless of the number of types of phones there exist, and they also hold if *x* and *y* are identical.

If we now let *T* be the set of all unique unordered pairs of phone architectures, we can formulate the tracing efficacy *E* of a centralized architecture as

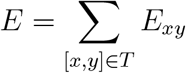

where

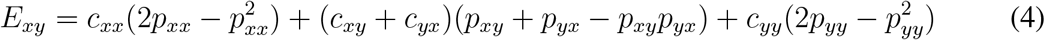

Here *c*_*z*_ denotes the probability that any given contact between two phones is of type *z* ∈ {*xx, yy, xy, yx*}. Note that in this formulation, *c*_*xy*_ = *c*_*yx*_, whereas *p*_*xy*_ and *p*_*yx*_ are distinct entities. The values for *c*_*z*_ can be calculated directly from the fraction of the different types of phones using the app. If we define *M*_*i*_ = *ϕ* as the proportion of apps running on iOS phones, and *M*_*a*_ = 1 *− ϕ* as the proportion running on Android, we have

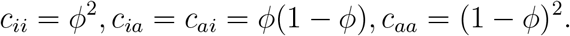

Note that the formulation in (4) mandates that data is collected centrally. If two phones are in proximity of each other, it suffices that the contact is detected by at least one of them. Note also that *E* is defined as the ratio of detected contacts to the total number of actual contacts *among app users*. In a situation with 100% app uptake in the population, *E* would be the total efficiency of the system in detecting contacts.

The mobile phone market in the world is dominated by two operating systems; iOS and Android, with significantly different properties. There is a rich set of different phone models as well, but the differences between the operating systems dominate the picture. We therefore instantiate our model by letting *T* = {[*i, i*], [*a, a*], [*i, a*]} where *i* means iOS, and *a* means Android. We used the aggregate contact dataset (see the Supplementary Information for details on the dataset) to calculate the number of detected contacts *D*_*ia*_, *D*_*ai*_, and *D*_*ia*+*ai*_, and from (2)-(3) we got the following probabilities:

- Probability that iOS detects iOS; *p*_*ii*_ = 0.54
- Probability that Android detects iOS; *p*_*ai*_ = 0.53
- Probability that iOS detects Android; *p*_*ia*_ = 0.53
- Probability that Android detects Android; *p*_*aa*_ = 0.74

More specifically, we focused on the most relevant contacts from an epidemiological point of view (i.e. within 2 meters and lasting at least 15 minutes)^18,33^. The numbers of detected contacts *D*_*ia*_, *D*_*ai*_, and *D*_*ia*+*ai*_ were computed directly since each contact is associated with a direction and labeled with phone types (i.e. phone *A* of type *x* detected phone *B* of type *y*).

In case of an app uptake less than 100%, we introduced a new set of contact probabilities 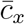, which are defined relative to the total population. Each value 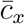 is the probability that a given contact occurs between two app users with the phone combination *x*, for *x* ∈ {*aa, ii, ai, ia*}. Above we had Σ_*x/*_ *c*_*x*_ = 1, since we only considered contacts between app users, but since a contact can also involve phones without the app installed, we have 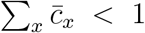, and the individual 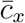 values depend on the uptake of the app among the respective users. We also assume that 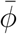 of the users have iOS devices and the remaining 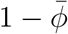 have android devices.

We then have

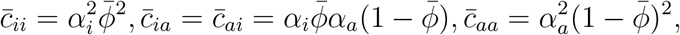

and as above the total efficiency is given by

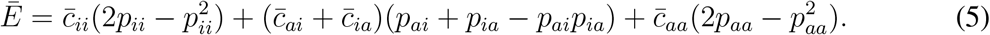

### Identifying random contacts

We used a random forest binary classifier^34,35^ to separate close contacts into known and random. To train a binary classifier, we needed a training set that includes both true positives (i.e. known contacts) and true negatives (i.e. random contacts). In absence of a verified ground truth, we needed to carefully pick these two sets from the underlying data. As true positives, we picked device pairs that met on at least seven different days. We examined contact patterns to discern potential true negatives. More specifically, we picked device pairs that were never in contact, despite both being in a repeated close contact with a common third device, as true negatives (see the Supplementary Information for more details).

We trained a random forest classifier with 20 trees, gini criterion, a maximum tree depth of 8, a minimum number of samples required to split an internal node of 2 and a minimum number of samples required to be at a leaf node of one. The values of these hyperparameters were selected after conducting an exhaustive grid search. Overall, we used nine features that were meant to capture the quality of information we have on a pair of devices, their connectivity as well as the topological commonalities between them (i.e. how many neighbours they share). We fitted three models for close contacts of any duration, at least five-minute long and at least fifteen-minute long. The three models exhibited an accuracy of 84%, 88% and 89%, respectively. We classified between 11% and 33.3% of contacts as random depending on the definition of close contact. Our model can classify contacts between devices with at least a single common neighbour. One-off contacts between devices without common neighbours could not be classified and were assumed to be known close contacts in order not to inflate the the added value of digital contact tracing. Hence, our estimates of the fraction random contacts are conservative. A detailed analysis of these aspects is provided in the Supplementary Information.

## Data Availability

The data and scripts for the efficacy model, the impact of uptake on pandemic progression and analysis of the ENS are available at the link below. Correspondence and requests for materials and datasets that are not yet public should be addressed to Ahmed Elmokashfi.

https://github.com/sundnes/smittestopp_data_model

## Supplementary Material

### 1 Smittestopp

As a response to a rising number in Covid-19 cases, the Norwegian health authorities decided on the 13th of March, 2020 to develop a contact tracing application. The app was subsequently launched on the 16th of April, which was received positively by the population^15^. The number of app downloads reached 1.5 million two weeks after the launch date. The number of active users per day (i.e. users that shared tracing information) peaked around 800k in the first few days post launch then decreased steadily to approximately 500k in early June (see Figure S1). The phone population was dominated by iOS devices but twice as many Android devices were lost in the course of the app deployment compared to iOS. The difference in the adoption rate between the two platforms reflects their popularity. The discrepancy in lost users can be blamed on the app resulting in a higher energy consumption on Android. Optimizing for Android was particularly hard given the high diversity in terms of both vendors and devices. The active users of Smittestopp were distributed across the country with a higher concentration in major urban hubs (see Figure 1b in the main text). Smittestopp was suspended on the 16th of June due to privacy concerns and decrease in infections^16^. Norway released a new app based on the exposure notification system in December 2020^17^. The source code of Smittestopp is publicly available^36^.

**Figure S1:**
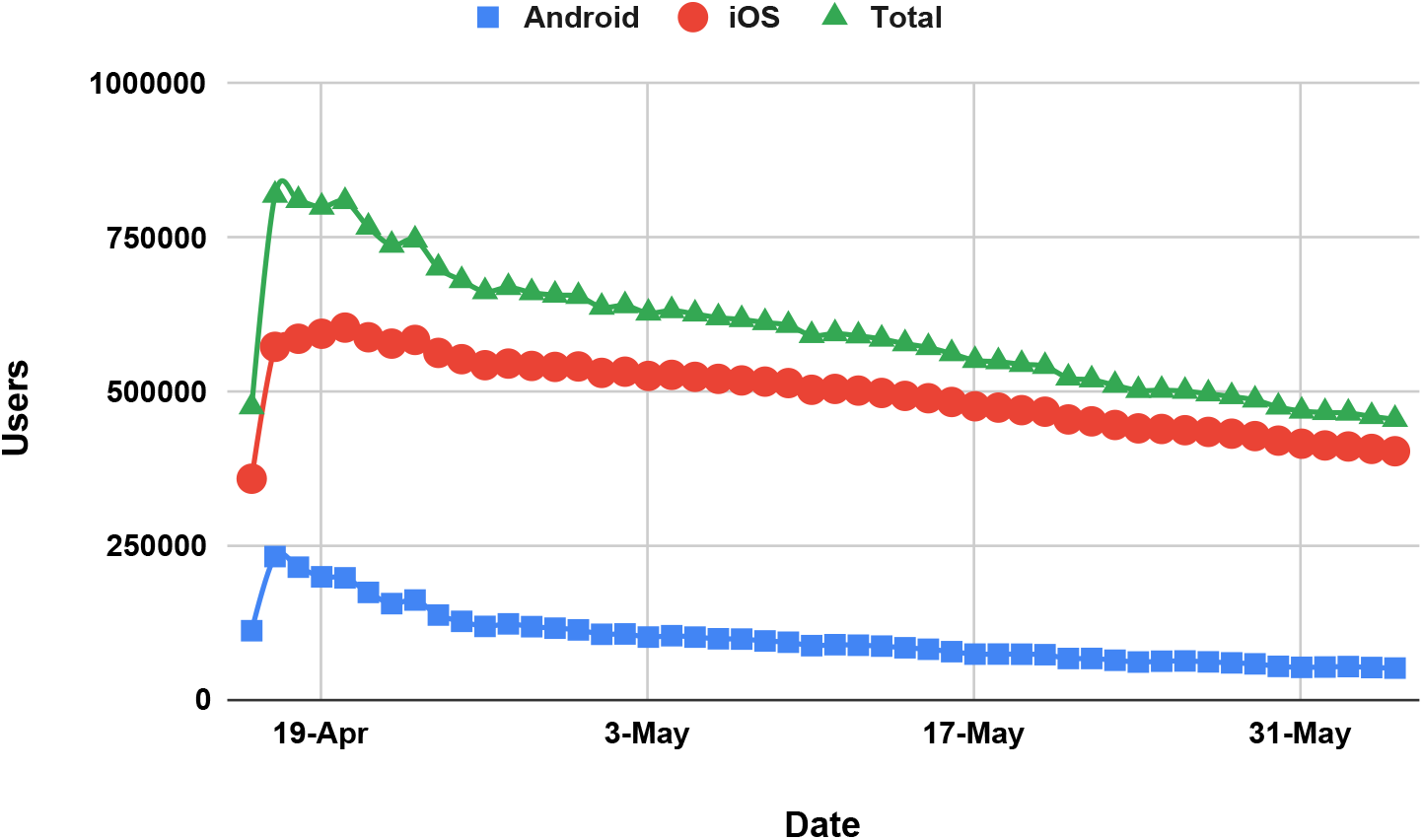
The number of active users per day.

Smittestopp was intended to both automate contact tracing and collect aggregate information on the mobility and interactions between users. This information was needed to help inform government policy on pandemic control. To this end, Smittestopp logged both users GPS locations and used Bluetooth Low Energy (BLE) to discover other users in proximity ^37^. Phones with Smittestopp would continuously broadcast their presence over BLE as well as periodically scan for phones with Smittestopp in proximity. Smittestopp used a universally unique service identifier for advertising presence and scanning for nearby devices. Note that BLE signals have a 10-meter propagation range. Upon the discovery of a nearby device, the phone would connect to it and measure the strength of the received BLE signal. Phone pairs acted independently meaning that the event a phone *A* discovering another phone *B* did not automatically translate to a discovery event in the opposite direction. In other words, the discovery in Smittestopp was asymmetric. Smittestopp was a centralized solution, that is all phones would upload their GPS and BLE measurments to a central database. This design choice was necessary for providing aggregate information on the mobility and interactions between users. It also transformed the discovery process from asymmetric to symmetric.

#### 1.1 The “iPhone-problem”

In the main text, we pointed to the fact that iOS and Android differ when it comes to the effectiveness of detecting nearby phones. The difference is related to limitations imposed by iOS on apps that run in the background, that is the app is running while the user is looking at another app or the screen is turned off. These limitations manifested in two forms. First, Apps using BLE and running in background were suspended by iOS a few second after entering the background mode. Suspended apps would still be visible to nearby phones that were active (e.g. Android phones or iOS devices with the app in the foreground). If a suspended app was discovered by another phone, iOS would bring it for approximately 30 seconds to the background, which means it could scan for other devices for a short while. Second, iOS would alter the format of BLE advertisement packets, sent by an app in the background, to a proprietary one. This makes it harder for other devices to discover nearby phones with Smittestopp running in the background^38,39^. These limitations implied that two iPhone devices with apps in background would not be able to detect each other. In other words, Smittestopp would not report that two iPhone users sitting next to each other in a bus with phones in their pockets as a close contact. This was a major hurdle given the large fraction of Smittestopp users that had iOS devices. Note that these limitations faced all apps that were not built using the Exposure Notification System^40^.

Smittestopp team eventually found a work around that would partially handle the aforementioned limitations. The work around leveraged the iBeacons and locations framework in iOS to periodically scan for BLE beacons in proximity^41,42^. A BLE beacon is a fixed device that periodically advertises its presence to nearby phones, which is typically used in indoor settings to help in navigation to various points of interest. Smittestopp scanned for non existent beacons, a positive side effect of this was that iOS would, during the scanning, relay BLE packets to apps in the background. The only caveat was that the screen needed to be on. So, a user flipping through the news, for example, was visible to nearby phones although the app was in the background. Even brief screen on events, for example in connection with the arrival of any notification, would make the phone detectable. The work around was rolled out in early May and immediately led to an increase in the detection rate for iOS devices (see Figure S2). The average number of detected devices increased steadily starting on the 8th of May. It continued climbing up as more users downloaded the update and eventually stabilized around the 18th of May (the red line in Figure S2).

**Figure S2:**
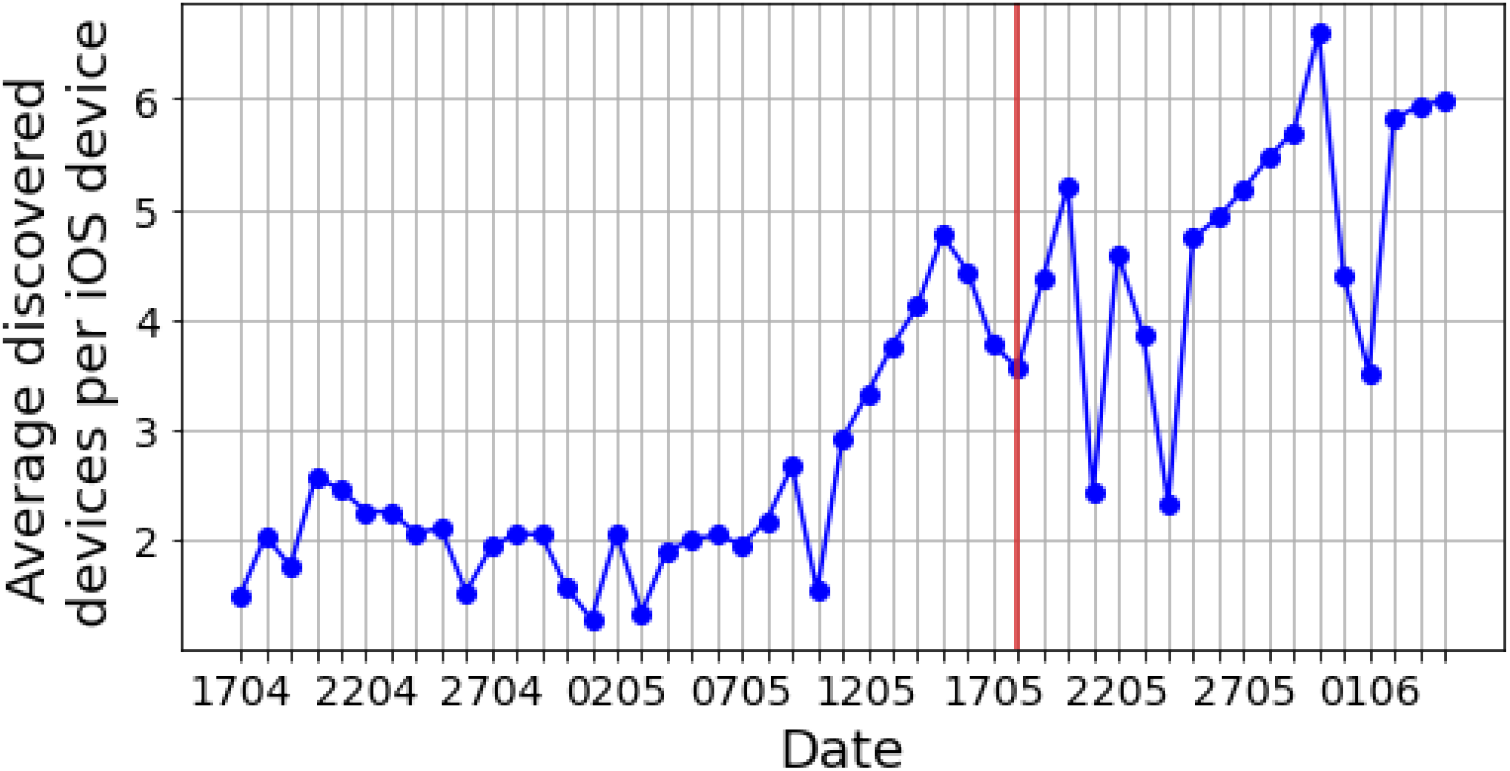
The average number of detected phone by an iOS device over time.

### 2 Dataset

Our dataset is an aggregated and anonymized version of the BLE measurements collected by Smittestopp. The original data was deleted, because Smittestopp’s privacy policy committed to deleting all raw data that was older than 30 days.

#### Data aggregation

The data was aggregated on daily basis. All device pairs that were in proximity were identified as contacts. A contact corresponds to a series of *N* BLE measurements, where *N* ≥ 1. Every measurement *i* is a tuple (*t*_*i*_, *RSSI*_*i*_), where *t*_*i*_ is the measurement times-tamp and *RSSI*_*i*_ is the measured BLE signal strength. All measurements in a contact, within a single day, were sorted, then the daily contact was defined as spanning the entire duration between the earliest and latest timestamps *t*_*i*_ and *t*_*N*_, respectively. Also the maximum and average RSSIs were recorded. As a result, a daily contact between two devices *p*_*a*_ and *p*_*b*_ is a tuple of (*p*_*a*_, *p*_*b*_, Δ*t, RSSI*_*max*_, *RSSI*_*avg*_, *N, T*_*a*_, *T*_*b*_), where Δ*t* is the time difference between *t*_*N*_ and *t*_*i*_, *RSSI*_*max*_ is the strongest signal strength, *RSSI*_*avg*_ is the average signal strength, *T*_*a*_ is the type of device *p*_*a*_ and *T*_*b*_ is the type of device *p*_*b*_. Note that *T*_*a*_ and *T*_*b*_ ∈ (*iOS, Android*). This approach to data aggregation masks all details about the time of contact, which is essential for ensuring users’ privacy. This has the side effect of mischaracterizing a pair of short encounters that were spaced by several hours as a single long varying contact. For example, two unrelated individuals that sat in the same train carriage in the morning and the afternoon of the same day. Filtering on the measured signal strength reduces the impact of such false positives by discarding encounters that are associated with a weak signal. Further, if such a contact was observed over a number of days with a signal strength that indicates a close spatial proximity, we can assume that the contact may qualify as a valid close contact.

#### Data anonymization

After aggregating encounters between device pairs into daily contacts, the device identifiers (i.e. *p*_*a*_ and *p*_*b*_ above) were hashed using the Secure Hash Algorithm 2 (SHA-2) with a 256 bits digest^43^. This algorithm produces a hash that can not be traced back to the original device identifier. It always, however, maps a device identifier to the same hash, which allows to track the activity of a device across several days. After the initial hashing step, each hash was mapped to a random number in the range (1, …, *N*), where *N* is the total number of devices, then the hashes were deleted. These random numbers were generated using the default seed which is the system clock timestamp. The mapping from the hashes to the random numbers is not reversible because the original hashes were deleted. Every contact in the dataset can not possibly be mapped to a particular individual, since the raw data was deleted. Further, the contact does not include additional spatial details like GPS coordinates or even any fine granular details on devices beyond being an iOS or Android device.

#### Basic contacts statistics

The dataset spans the period from the 17th of April to the 4th of June 2020, that is from the first day after Smittestopp was released to ten days before it was suspended. As explained in Sec .1.1, Smittestopp underwent a major update in early May to address the iOS-imposed limitations. This translated into a higher rate of false negatives in the first three weeks of Smittestopp’s lifetime. We therefore considered mainly the data collected between the 18th of May and 4th of June, that is a total of 18 days. In this 18-day period no updates of the app were pushed to the phones, and the effects of previous updates of the app had stabilized. In order to avoid spurious users who downloaded the app and stopped using it after a short while, we removed from the dataset all devices that were not present on seven different days (i.e. the device was seen in a contact on seven different days). We also considered only contacts between devices that had a 7-day overlap, meaning that both devices appeared in the dataset on at least the same seven days.

We further derived three datasets as follows:

1. *Proximity Contacts (PC)*. These are contacts of any duration but with an average RSSI that is consistent with 2 meters proximity. To convert from RSSI to distance, we used the thresholds that were set by Smittestopp^22^, where contacts with *RSSI* ≥ −85 dBm were considered as close.
2. *Relatively Risky Contacts (RRC)*. These are contacts that were at least 5 minutes long with an average RSSI that is consistent with 2 meters proximity.
3. *Potentially Risky Contacts (PRC)*. These are contacts that were at least 15 minutes long with an average RSSI that is consistent with 2 meters proximity. PRC contacts are associated with a higher covid-19 transmission risk, if one of the involved individuals is contagious.

Accordingly, *PRC* ⊂ *RRC* ⊂ *PC*. Table. S1 summarizes the number of contacts and devices in our dataset. The number of unique devices is about 545k, which amounts to 12.5% of the Norwegian population over 16 years old. ^1^ The number of unique devices drops as the dataset becomes stricter. Overall, 10.8% of all contacts were *PRC*.

**Table S1:** Basic statistics of contacts

|  | All | PC | RRC | PRC |
| --- | --- | --- | --- | --- |
| Unique devices | 545354 | 525646 | 485730 | 461068 |
| Unique contacts | 26792433 | 10665399 | 3946986 | 2889922 |

### 3 Bluetooth and False Positives

Our approach for estimating the efficacy of Smittestopp does not capture the number of false positives generated by the system. We do not believe that this information is to be found in the dataset, as the dataset contains no ground truths on proximity. These false positives stem from the fact that the BLE signal can exhibit non-trivial propagation patterns depending on surrounding environments^5^. For example, environments with metallic elements like the inside of a tram can amplify BLE signal and thus underestimate the actual distance. Also phones separated by thin walls may appear closer than they really are as far as the virus spreading is concerned.

We, however, argue that the number of false positives generated by BLE is at the level of noise when compared to the false positives stemming from accidental lack of infection spread. Let us first define a false positive contributed by the technology to be a registered contact, where the definitions of what constitutes a contact does not hold. Since BLE will only communicate over short distances, usually up to 10 meters, a false positive according to this definition will amount to situations where the following holds:

- There is a proximity between the two persons of 10 meters or less. If this was not the case the phones would be out of range for each other’s BLE signal, and no registration would be made.
- The proximity between the persons lasted for 15 minutes or more - otherwise it would not be registered as a contact.
- The true distance between the two persons would be more than 2 meters - otherwise it would not be a false positive.
- The measurements from BLE falsely indicate a distance of less than 2 meters for 15 minutes.

There will be situations in daily life where these four requirements hold, for example people seated within 10 meters of each other on a bus or in a theater. Still, for this to be a dominating factor, two things must be true. First, the disease must be extremely contagious, so that most contacts within 2 meters in 15 minutes get infected. Second it must be extremely accurate so that contagion stops at 2 meters, and starts after 15 minutes. This is far from being the case for Covid-19. Various studies estimate the secondary attack rate for Covid-19 at 17% for household contacts and 27.8% to contacts who were spouses of index cases^44,45^. The secondary attack rate is markedly lower for settings outside the household^20,46^. Accordingly, both manual and digital contact tracing will yield a high fraction of false positives from an epidemiological point view, that is identified contacts that did not contract the virus.

### 4 Classifying contacts

Contacts can either be known or random. Manual contact tracing can in principle identify all known contacts given that the case recalls all recent encounters. Random contacts, however, can not be identified by manual contact tracing alone. An additional approach that tracks the presence of unrelated individuals in a particular location at a particular time, like digital contact tracing or restaurants guest lists, is needed.

Our dataset does not include any extra information, like GPS coordinates or a user-provided context, to help separating known from random contacts. Nevertheless, it tracks contacts over time as well as contact duration and these two can provide an idea about repeated long encounters. The contact dataset can also be represented as a network with devices as nodes. A pair of nodes are connected, if a contact is recorded between them. The corresponding edge weight is the number of unique days with contacts. This contact graph can give insights into similarity between devices, in terms of presence of common neighbours, which can be used in inferring known contacts.

The lack of ground truth, however, complicates the task of validating the outcome of the classification process. To address this, we leveraged the intuition that random encounters tend to be shorter than known contacts as well as unlikely to repeat. In addition, since the data was collected as the first wave of the pandemic was receding, human mobility was still limited and the society was in many ways closed, we would expect a relatively small number of random encounters.

#### One-off contacts

A plausible starting point, when identifying random contacts, is to look at one-off contacts, i.e., contacts that were observed on only one day in our dataset. This one-off behaviour was observed despite the fact that the involved devices were simultaneously present, that is they have registered contacts with other devices, in at least seven days. In other words, we can not explain the lack of several contacts by simply non-overlapping activity periods.

We now focus only on the three derived datasets and ignore the raw contacts since these involve many contacts that are associated with distances greater than two meters. Table S2 shows the percentage of one-off contacts for each dataset. The percentage of one-off contacts decreased as we tightened the contact selection criteria. This is expected since the tighter the criteria the more likely we avoid spurious contacts. Given the state of the society at the time, we would expect a lower extent of random contacts. Accordingly, these numbers are likely to involve known close contacts, since these also were recommended to social distance. For example, friends and family who only met once during the study period. Other causes like app-related artefacts; users tendency to switch off the app when they are home and only use it when outdoors or simply people living in larger homes end up leaving their phones in separate rooms which results in a lower signal strength might have also contributed to the high fraction of one-off contacts. Hence, the percentage of random contacts will likely be greatly overestimated if we assume that all one-off contacts were random contacts.

**Table S2:** One-off contacts.

| **Dataset** | **% of one-off contacts** |
| --- | --- |
| PC | 77.1% |
| RRC | 64.1% |
| PRC | 57.3% |

#### Simple filtering

A simple approach would classify an one-off link between two devices *a* and *b* as random if and only if there exists no third node *c* that is connected to both *a* and *b* (i.e. *a* and *b* do not have common neighbours). This classifier, which we refer to as *naive* in the following, can result in false negatives by classifying an actual random link as non-random. For instance, two devices, which are actual close contacts, encountered a third random contact while traveling together. These devices will form a triad that would be interpreted as a sign of a non-random contact. It can also lead to false positives by classifying known contacts as random contacts. For example, friends that met only once during the study period without being in proximity to a third person simultaneously. Another naive approach for identifying random contacts is to impose a minimum time duration for accepting a contact as non-random, we refer to this classifier in the following as *time-limited*. Here, we do not take into account whether the contacts had a common neighbor or not. We set this threshold to 60 minutes, which should be enough to capture encounters with friends and neighbours for example. While this may seem plausible, such a minimum duration is a function of the type of encounter as well as the app and can vary widely.

Table S3 shows the percentage of one-off contacts after applying the *naive* and *time-limited* filtering. The two approaches reduced the percentage of one-off contacts but with different magnitudes.

**Table S3:**
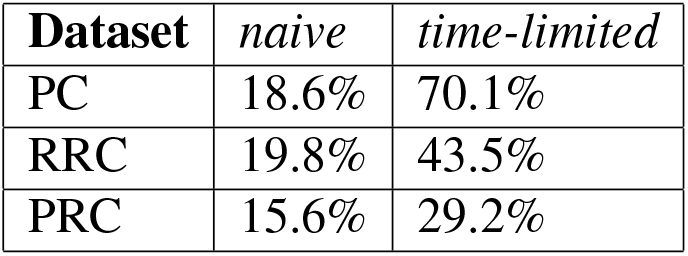
Percentage of one-off contacts after applying simple filters.

| **Dataset** | *naive* | *time-limited* |
| --- | --- | --- |
| PC | 18.6% | 70.1% |
| RRC | 19.8% | 43.5% |
| PRC | 15.6% | 29.2% |

The *naive* approach returned comparable percentages of one-off contacts for the three datasets. This is unexpected given the underlying differences between the datasets with respect to minimum contact duration. The number of one-off contacts is expected to be the highest for PC and lowest for PRC. The *naive* approach is simply indicating that the likelihood of having no common neighbor is invariant to the contact duration and is consistent across the three dataset. The *time-limited* approach returned higher percentages of one-off contacts, which means that the majority of the original one-off contacts lasted less than an hour. Here, the three dataset exhibited differences that are consistent with the underlying differences between them. We further looked at the duration of the one-off contacts that were flagged by the *naive* approach in table S3 (i.e. those between users without common neighbours) and those which were not flagged. Table S4 shows the percentage of contacts that were longer than one hour for both categories. Contacts between users with common neighbours were more likely to be longer. The flagged contacts involved a non-trivial percentage of long contacts, especially for PRC, which hints at the presence of false positives. The not flagged contacts included also a large fraction of short contacts, which suggests that the *naive* approach had failed in identifying a sizable fraction of random contacts (i.e. false negatives).

**Table S4:** Percentage of contacts longer than one hour for one-off contacts that were flagged (left as one-off) and not flagged by the naive approach.

| **Dataset** | *Flagged* | *Not flagged* |
| --- | --- | --- |
| PC | 3.0% | 11% |
| RRC | 15.6% | 39.5% |
| PRC | 30.9% | 55.7% |

In summary, the two approaches yielded different results. The *naive* approach did not account for the underlying differences between the three datasets. This resulted in inferring close estimates of potential random contacts and apparently sizable fractions of both false positives and false negatives. The results of the *time-limited* approach were compatible with differences between the underlying datasets, which is expected as these differences are indirectly captured by the time threshold. However, this approach does not take into account the structure of the contact graph, which would make it vulnerable to false positives. Accordingly, a successful approach for identifying random contacts must yield results, with respect to the volume and duration of random contacts, that are consistent with the differences between the underlying dataset. Further, the results should indicate clear qualitative differences between known and random close contacts.

#### Machine learning classifier

To overcome these limitations, we employed a random forest binary classifier that takes into account a broader set of features^34,35^. Using a supervised classifier, we aimed to identify relationships between the features beyond the presence of common neighbours as well as to avoid imposing arbitrary thresholds on contact duration.

To train a binary classifier, we needed a training set that includes both true positives (i.e. close contacts) and true negatives (i.e. random contacts). In absence of a verified ground truth, we needed to carefully pick these two sets from the underlying data. Intuitively, *true positives* will be users that have a high frequency of daily encounters, for instance people living or working together. We accordingly picked device pairs that had at least seven encounters, that is they met on seven different days. Identifying *true negatives* is more challenging though. One could think of picking random pairs of devices with no common neighbours. This can be a viable approach, if all devices occupy the same physical space and they can plausibly meet. However, our data covers the whole of Norway, which reduces this plausibility and renders the above approach inadequate. We instead reverted to the contact pattern to discern potential true negatives. Assume that devices *a* and *b* have met each others frequently, while device *c* has more than two encounters with device *a* but has never been in the proximity of device *b*. The higher the frequency at which *a* and *b* meet, the less likely that *c* has ever been in the proximity of *b*. In this example, *a* and *b* could be family members and *a* and *c* coworkers, so *b* and *c* are true negatives that never met during the data collection period despite the presence of common neighbours. Note that we did not require that *a* and *c* have only one common neighbour, since that would directly influence the definition of random contacts. We expected nevertheless true negatives to share fewer neighbours if any, which serves as a reasonable test to verify the identified true negatives. To confirm this, we plotted the density of the number of common neighbours for both true positives (TP) and true negatives (TN) (see Figure S3). The plot matched our expectations. TNs shared only one neighbour in most cases.

**Figure S3:**
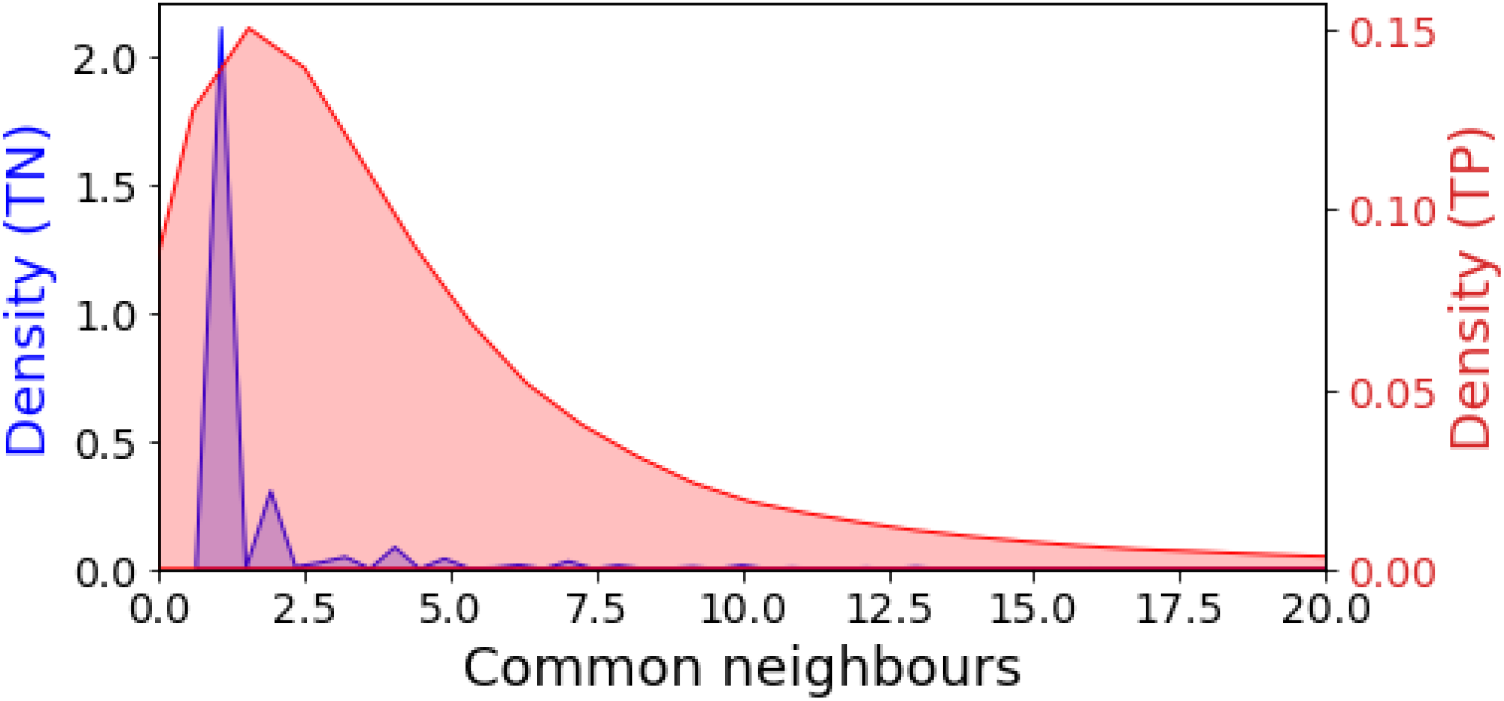
The density of the number of common neighbours for both the inferred true positives and true negatives. TNs have clearly fewer common neighbours.

##### Features

We trained our model using a total of nine features, which were meant to capture the quality of information we have on a pair of devices, their connectivity as well as the topological commonalities between them.

1. device visibility and overlap features: for a pair of devices *a* and *b*, we collected the number of days each device was active, i.e., either discovered or was discovered by another device. We also collected the number of days both devices were active, which we refer to as availability overlap. These features were meant to control for the effect of devices’ measurement coverage on the likelihood they discover each other repeatedly.
2. Graph and topological commonalities features: for a pair of devices, we collected each device degree (i.e. the total number of unique contacts it recorded) and the number of common neighbors. We further computed two features to capture the similarity in connectivity: Jaccard’s index ^47^ and Adamic/adar (AA) index^48^. For a pair of devices with a set of neighbours *N* (*a*) and *N* (*b*), Jaccard’s index is given by 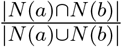, which is basically the fraction of common neighbours. The AA index is defined as the summation of the inverse logarithmic degree centrality of the neighbours shared by pair, it is given by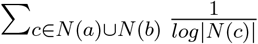. Essentially, the AA index is higher if devices tend to commonly connect to low degree devices than high degree ones. Both metrics have been used before to predict missing links in social networks^49^.
3. device type features: for each pair of devices, we input their device type(s), which is either Android or iOS.

Note that the aforementioned features do not include contact duration since we do not have a clear mapping between contact duration and type. Also, these features can only help classifying one-off contacts with common neighbours, that is the difference between the initial one-off contacts in Table S2 and those identified by the *naive* approach in Table S3. For example, it will classify over 70% of the one-off contacts in the PRC dataset. Classifying one-off contacts without common neighbours requires extra features about contexts of contacts that do not exist in our dataset. We argue that this is not a major limitation because the majority of one-off contacts were between users with common neighbours. More specifically, the share of one-off contacts between users with common neighbours is 75.8%, 69.2 and 72.8% for PC, RRC and PRC respectively. Also assuming that one-off contacts without common neighbours are not random will give a conservative estimate of the share of random contacts.

##### Classifier

We train a random forest classifier with 20 trees, gini criterion, a maximum tree depth of 8, a minimum number of samples required to split an internal node of 2 and a minimum number of samples required to be at a leaf node of one. The values of these hyperparameters were selected after conducting an exhaustive grid search.

##### classification accuracy

We fitted three models for the PC, RRC and PRC dataset, respectively. We performed k-fold cross validation for each model to verify that our models can generalize. The three models exhibited a high level of accuracy as follows: PC (84%), RRC (88%) and PRC (89%). Tables S5, S6 and S7 present the confusion matrices for the three datasets.

**Table S5:** PC confusion matrix.

| Real<br>contact \ Predicted<br>contact | Random | Close |
| --- | --- | --- |
|  | Random | Close |
| Random | 80.5% | 19.5% |
| Close | 12.2% | 87.8% |

**Table S6:** RRC confusion matrix.

| Real<br>contact \ Predicted<br>contact | Random | Close |
| --- | --- | --- |
|  | Random | Close |
| Random | 86.0% | 14.0% |
| Close | 9.6% | 90.4% |

**Table S7:** PRC confusion matrix.

| Real<br>contact \ Predicted<br>contact | Random | Close |
| --- | --- | --- |
|  | Random | Close |
| Random | 86.7% | 13.3% |
| Close | 8.3% | 91.7% |

All three models exhibited a higher accuracy when classifying close contacts. The performance, however, slightly degraded when classifying random contacts, where a higher fraction of them was classified as close contacts. Hence, these models are conservative when it comes to flagging an encounter as a random contact. This is a desirable property, because this way the model will not lead to overestimating the benefits of digital contact tracing. We also note that the accuracy of the model improved as the definition of contacts became stricter. The stricter the definition the less likely the contact is a false positive. Consequently, classifying these contacts will be less error prone.

##### Feature importance

We also investigated the role of different features and their contribution to the model’s accuracy. To this end, we use the Gini importance or the Mean Decrease in Impurity measure, which counts the fraction of times a feature is used in determining how to split the classification tree.

Table S8 shows the Gini’s importance for all features. AA’s index and the number of common numbers were important to more than half of the decisions. Then followed by the features that describe the quality of the measurement data. Accordingly, the model has learned to classify contacts depending mainly on the features that capture topological commonalities between devices. Note that although the number of common neighbours and Jaccard’s index capture some of the topological similarity aspects that are captured by AA-index, the latter is more important for discriminating contacts. This could be attributed to the fact that AA-index refines the neighbourhood comparison beyond simple counting by considering structural similarities.

**Table S8:** Features importance.

| **Feature** | **Gini importance** |
| --- | --- |
| AA's index | 0.398 |
| Number of common neighbours | 0.162 |
| Device A total days with measurements | 0.122 |
| Device B total days with measurements | 0.116 |
| Jaccard's index | 0.102 |
| Total overlapping days with measurements | 0.079 |
| Device A's degree | 0.007 |
| Device B's degree | 0.007 |
| Type of phones | 0.007 |

#### Inferred random contacts

The random forest model classifies a non-trivial fraction of oneoff contacts as close contacts, which we summarize in Table S9. Only 11% of PRC contacts were classified as random. If we consider only the contacts that we could classify (i.e. ignore one-off contacts between pairs without common neighbours), the percentage of random contacts increased slightly to 40.7%, 18.3% and 13.1% for PC, RRC and PRC, respectively. The inferred fractions of random contacts are similar to numbers suggested by previous studies of social contacts as well as reports on untraceable Covid-19 infections^19,50^. Figures S4 and S5 show how the fraction of random contacts has evolved in period for the PC and RRC datasets, respectively. Our estimates of random contacts are conservative, since all one-off contacts between users without common neighbours are assumed not to be random contacts. Now, if we assume that all such contacts lasting less than an hour were random, the fraction of random contacts would increase to 20.8%, 31.2% and 51.4% for PRC, RRC and PC, respectively.

**Table S9:** Percentage of one-off contacts after applying the random forest classifier.

| Dataset | Random contacts (%) |
| --- | --- |
| PC | 33.3% |
| RRC | 14.5% |
| PRC | 11.0% |

**Figure S4:**
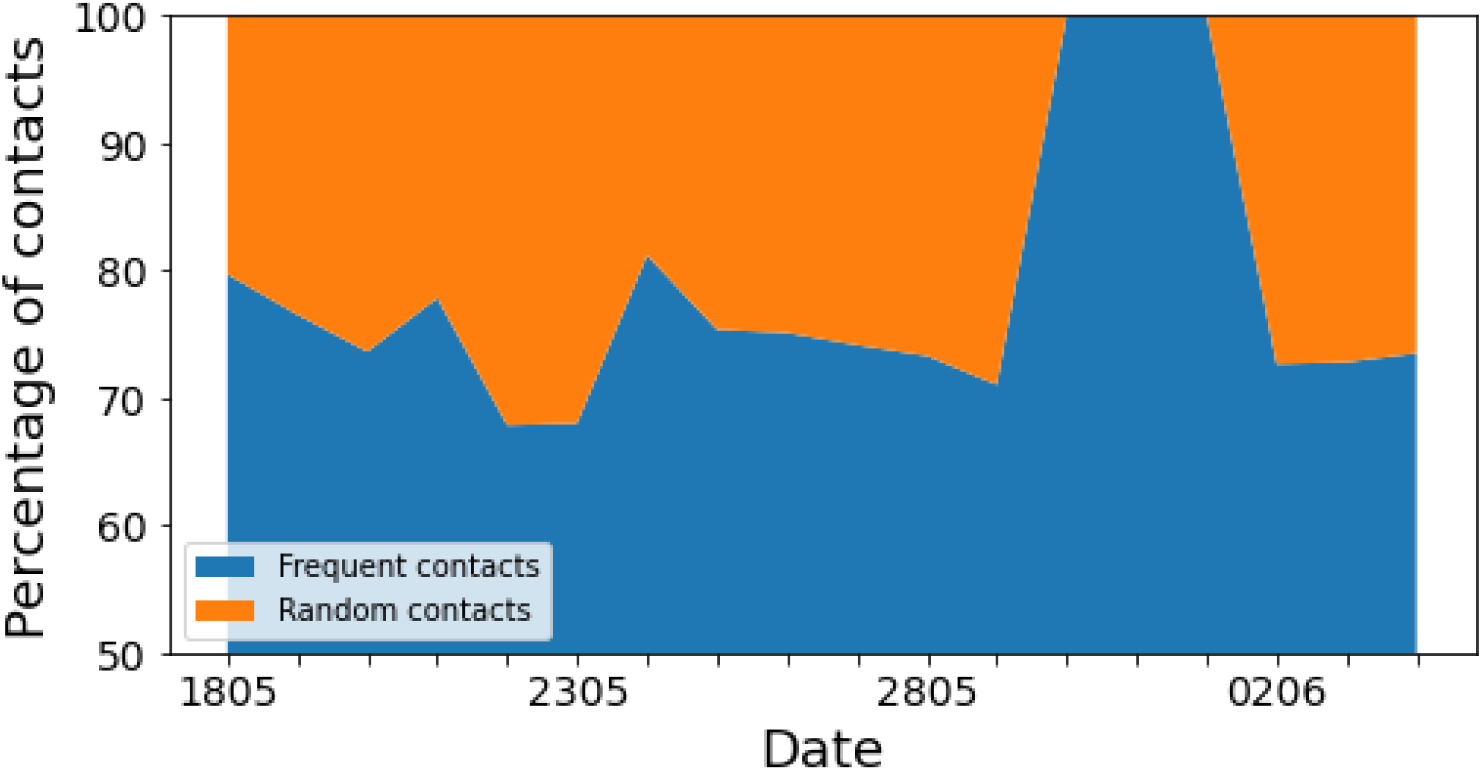
The timeseries of the fraction of random contacts for the PC dataset.

A limitation of our approach is that the underlying contact graph is incomplete, that it does not include the entire adult population of Norway. This incompleteness can lead to classifying close contacts as random contacts. We attempted to minimize the impact of this by including users that sent data on at least seven days as well as pair of devices that were sending data on at least the same seven days. We also considered all one-off contacts between users without common neighbours as close since we did not have features that captured the contexts of these contacts. We could not gauge the impact of graph incompleteness on our inference because we lacked ground truth. However, the qualitative differences between random contacts and close contacts in terms of contact duration and number of contacts of each type (see Figure 3 in the main text), suggest that we are discriminating contacts of different underlying characteristics.

### 5 The effectiveness of exposure notification system-based contact tracing apps

The exposure notification system (ENS) developed by Apple and Google has emerged as the de-facto standard for digital contact tracing^51^. Currently, 28 countries and 19 US states US have already rolled out ENS-based apps^52^. Key to the success of the ENS is the built-in privacy preservation and expected superior performance.

The built-in privacy preservation, however, makes the task of precisely assessing apps effectiveness impossible. If an app user tested positive for Covid-19, he or she is handed a one-time code to register the test results in the app, which in turn uploads a set of keys, one key per day for at most the last 14 days, that identifies the index case to a central server. Other app users download periodically all uploaded keys and match them with all saved encounters to determine whether they have been in proximity of an index case. In this process, health authorities hand out the one-time codes and configure the ENS to define close contacts.

**Figure S5:**
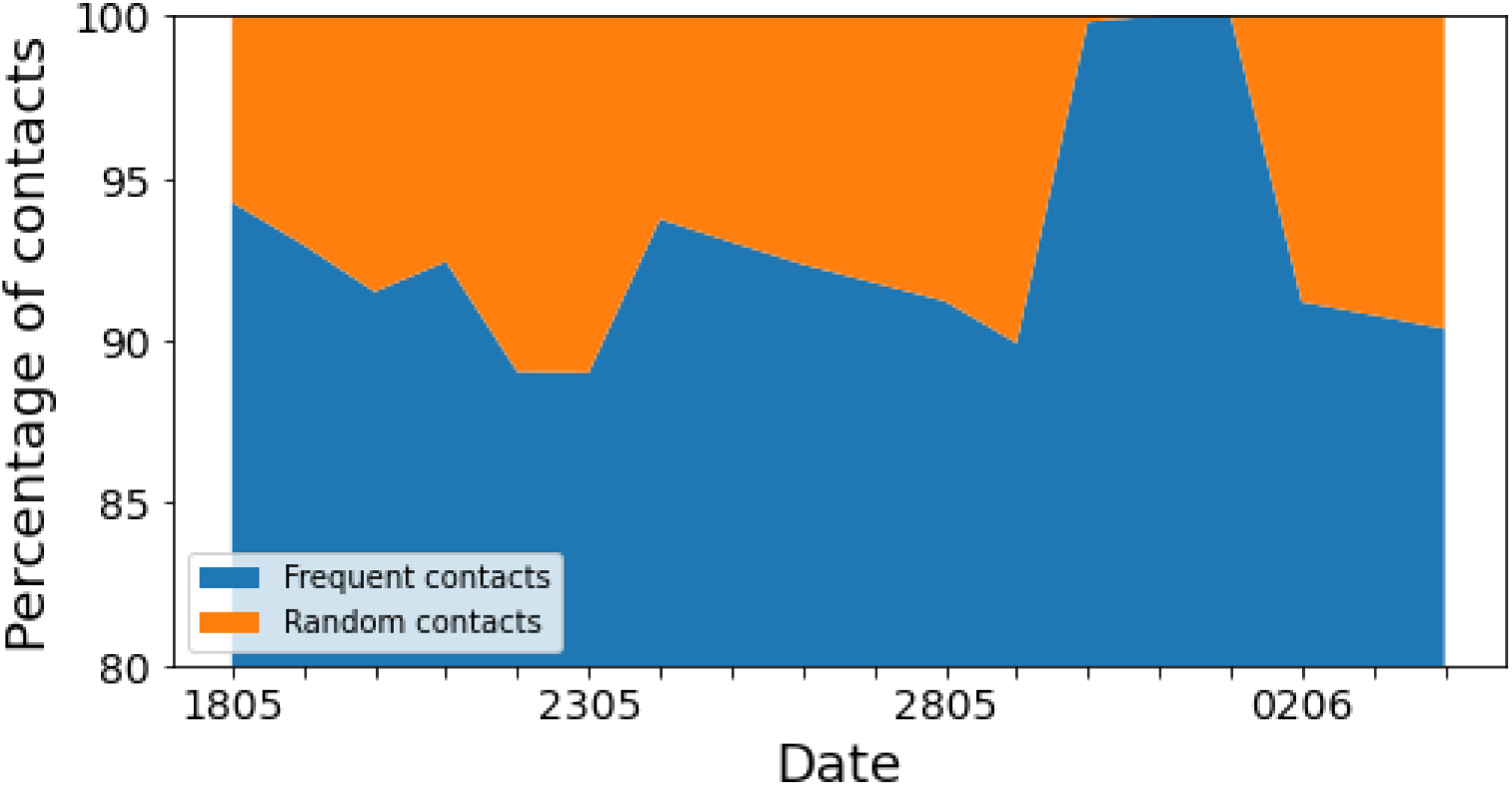
The timeseries of the fraction of random contacts for the RRC dataset.

Quantifying the effectiveness of ENS-based apps is a multi-step process. The first step verifies the accuracy of inferred contacts. The accuracy depends on the suitability of BLE signal attenuation in inferring distances and the configuration of the ENS. Several efforts have examined these two aspects and showed that ENS, like any other BLE-based distance estimation system, can have unpredictable performance due to wireless propagation artefacts^4,22^. They also showed that ENS configurations in use by many countries tend to miss real contacts. As a response many countries are continuously monitoring their configurations. Note that all these efforts were limited to a small number of phones, usually less than hundred. The second step tracks the adoption rate of the app, that is the fraction of population that have installed the app, as well as the fraction of index cases that have the app installed. Unfortunately, both numbers can be at best approximated. Health authorities can collect statistics from app stores about how many users have downloaded the app, but these app stores do not track uninstalls. They can also instrument the apps, by for example asking each app to connect to a central server to check for configuration changes *x* times per day^53^. There is no automated way to check whether an index case has the app. Manual contact tracers or laboratories issue the case a one-time code to report the diagnosis to the app, which in turn triggers the upload of exposure keys. This has inherent limitations since the case can choose not to report having the app. Furthermore, the index case can also choose not to report the diagnosis. Health authorities can track both numbers since they issue the one-time codes and later verify them. The last step measures the epidemiological benefit by tracking false positives and false negatives in comparison with manual contact tracing. These can be estimated by surveying individuals tested for Covid-19 about the use of the app and whether they were notified through it. The test results can then reveal whether digital contact tracing is identifying epidemiologically risky contacts.

The available followup data on deployed ENS-based apps is limited. We used publicly available statistics about the German (Corona Warn) and Swiss (SwissCovid) apps to gauge their effectiveness^24,25^. These two apps were rolled out in June 2020. In particular, we used the published numbers about diagnosis upload via the app, the app downloads and the number of inquiries to the health system following the reception of an exposure notification. We analyzed 3 months worth of data for Corona Warn and over 4 months for SwissCovid.

Figures S6 and S7 present three key measures for both apps. The app coverage, that the fraction of index cases with the app installed and have requested a one-time code, follows closely the apps uptake ratio. Further, the coverage remains at the same level as the number of cases surges. Hence, the app uptake seems to be a good proxy for gauging the case coverage. Each uploaded one-time code generates at least one call to the app hotline, a phone number that is only revealed when a user is notified of a potential risky contact, which is an indirect indicator of the effect of the app. Note that Corona Warn makes available both the number of people received one-time codes and those decided to enter them, while SwissCovid makes only the latter available. Between September 2020 and January 2021, only 55% of index cases uploaded the received one-time codes to Corona Warn. Earlier analysis reported a higher fraction, 66.2%, for SwissCovid^54^. Both numbers are low essentially halving the effectiveness of the apps.

**Figure S6:**
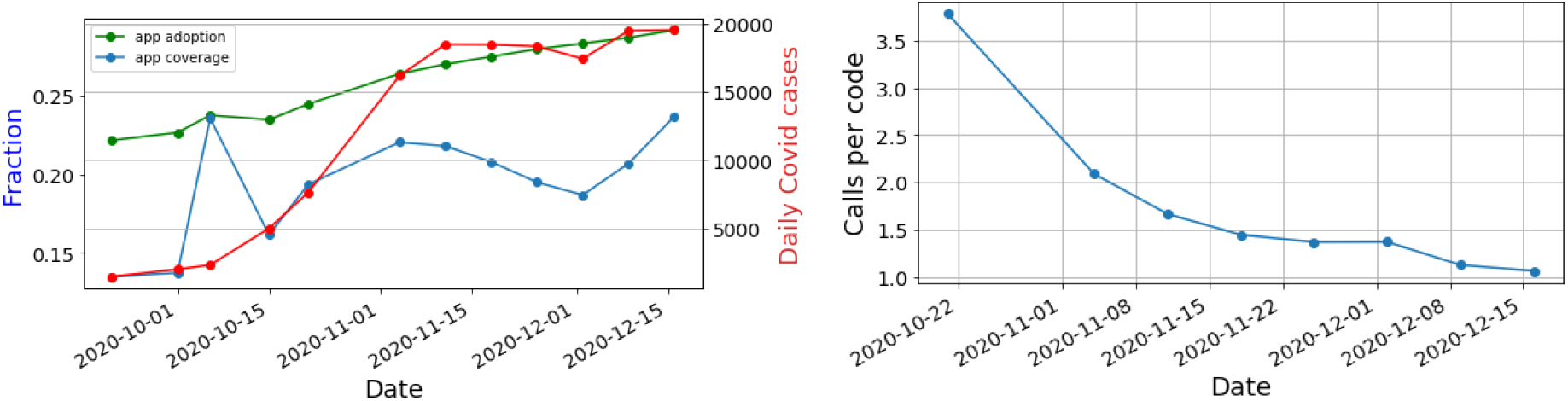
(**Left**)Left Y-axis: the adoption rate of Corona Warn measured as the fraction of the German population that downloaded the app and the app coverage measured as the fraction of index cases that requested one-time codes. The right Y-axis shows the daily confirmed Covid cases. (**Right**) the average number of calls to the Corona Warn hotline per each issued one-time code

**Figure S7:**
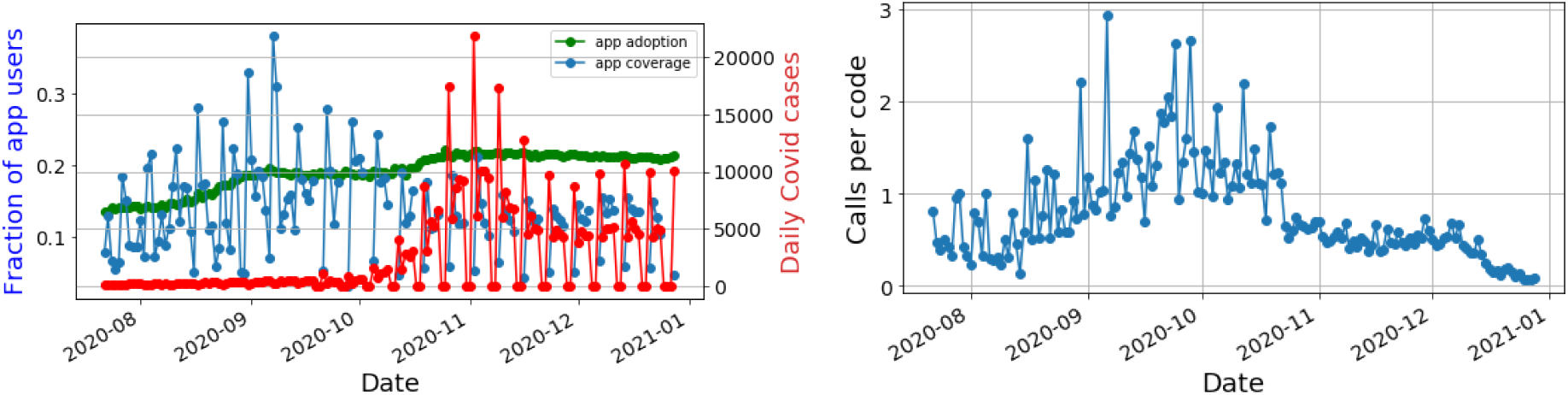
(**Left**)Left Y-axis: the adoption rate of SwissCovid measured as the fraction of the German population that downloaded the app and the app coverage measured as the fraction of index cases that requested one-time codes. The right Y-axis shows the daily confirmed Covid cases. (**Right**) the average number of calls to the SwissCovid hotline per each issued one-time code

We also looked at average app coverage in comparison with adoption rate for Denmark based on the publicly available numbers^55^. The Danish app was rolled out in June and it has been, by January 3rd 2021, downloaded by 35% of the population. The average coverage of the Danish app is at 29% compared to 14% and 11% for Switzerland and Germany, respectively. About 80% of index cases in Denmark, with app installed, have opted to submit their test diagnosis. Denmark goes one step further by reporting the number of individuals that submit to testing following an app notification and the corresponding true positive rate for the PCR test. This rate is at 0.9% for app users compared to 4.2% for all tested individuals. While this rate may appear low, recent epidemiological findings show that the infection rate is far lower for random/casual contacts, which is exactly the type of contacts that digital contact tracing aims to find. More specifically, Cheng et al. showed that, in Taiwan, family contacts had a secondary attack rate between 4.7% and 5.3% compared to 0.9% and 0.1% for health personnel and other less frequent contacts^20^.

### 6 Estimating the efficacy of manual contact tracing

An attempt to estimate the number of traceable contacts is provided by Kucharski et al.^56^. The study is based on the BBC Pandemic dataset, which contains self-reported contacts in the form of a face-to-face conversation (*>* 3 words) or a contact involving physical touch. Contacts were grouped into household contacts, work, school, and other. For each contact, responders also reported whether they knew this person (i.e. if had they met before). Kucharski et al. classified a contact as non-traceable if the two persons had not met before, and arrived at the following proportion of traceable contacts: household contacts 100%, school contacts 90%, work 71%, and other contacts 52%. These proportions were combined with the mean number of contacts in each category, listed in Table S10 to estimate the total number of traceable and non-traceable contacts. For instance, a person over 18 will have, on average, 7.62 traceable contacts and 5.26 non-traceable contacts per day.

**Table S10:** Mean number of contacts in each catoegory, calculated from the supplementary data provided with^56^.

|  |  |  |  |
| --- | --- | --- | --- |
| Under 18 |  | Over 18 |  |
| Household | 3.02 | Household | 1.90 |
| School | 5.54 | Work | 5.94 |
| Other | 1.47 | Other | 3.74 |

There are several limitations in the use of the BBC pandemic dataset for estimating traceable contacts. Since the data is based on self-reporting, it only considers contacts in the form of conversations and physical contact. Other contacts, such as being in close proximity on public transportation, are not included. Furthermore, there is (to our knowledge) no threshold on the duration of a contact. A very short exchange of words, lasting less than a minute, will still be counted as a contact, while manual contact tracing typically uses ten minutes as a guideline threshold.

Authorities in many countries publish reports on infected cases and the source of infection, which provides an alternative data source for estimating the efficacy of manual contact tracing. For instance the local health authorities in Oslo, Norway reported 423 cases from October 19 to October 25, 2020. For 96 of these cases, (23%), the source and location of infection were not known. From October 26 to November 2, 179 (25%) out of 716 reported cases had an unknown source^50^.

#### The impact of tracing random contacts on the pandemic

If we assume, for instance, 60% app uptake in the population, we observe from Figure 2 in the main text that the efficacy of the app tracing is approximately 30%. We may further assume that the app is used as a supplement to manual contact tracing, and that its main purpose is to trace contacts that are not traceable by manual tracing. Based on the BBC pandemic dataset, Kucharski et al^56^ estimated that 41% of contacts were non-traceable. This estimate gives maximum efficacy of 59% and adding digital tracing with 30% efficacy will increase the overall tracing efficacy to 71%. As previously demonstrated by Ferretti et al^3^, a difference of this magnitude can easily mean the difference between a controlled pandemic and an exponential growth of cases.

### 7 Modeling the effect on the pandemic spread

We have used the model of Ferretti et al^3^ to quantify the potential effect of the digital contact tracing on the pandemic spread. In^3^ the model was used to quantify the effect of efficacy and delay in isolation of infected individuals and tracing of their contacts, while for the present study our primary interest is the potential effect of app uptake. To quantify this effect, we calculated the tracing efficacy as a function of app uptake for the two operating systems, given by Eq. 5 in the main text, and used these numbers as input to the model. Although efficacy of self-isolation and delays in quarantining and isolation are not a a primary focus of the present work, we used multiple values for these parameters to investigate their potential impact on results and conclusions. The model parameters that were varied in our calculations are specified in Table S11. All other parameters were fixed at their default values specified in^3^.

**Table S11:** Parameters that are varied in our application of the model from^3^ to assess the effect of app uptake on the pandemic spread.

| Parameter | Value(s) | Source |
| --- | --- | --- |
| Reproduction number ($R_0$) | 1.5, 2.7 | <sup>57</sup> |
| Efficacy of contact tracing | 0-100% | Calculated from Eq. 5 in the main text |
| Efficacy of case isolation | 50%, 70%, 90% | <sup>3, 56, 58, 59</sup> |
| Delay of tracing and isolation | 4 hrs, 24 hrs, 48 hrs | <sup>3</sup> |

Figure S8 shows the growth rate as a function of app uptake for *R*0 = 1.5, for case isolation efficacy of 50% (top), 70% (middle) and 90% (bottom), and for delays of 4, 24, and 48 hours, respectively, from left to right. The black line shows the limit *r* = 0, which marks the difference between exponential growth and a decaying pandemic. As previously demonstrated in^3^, both the delay and isolation efficacy impact the overall effectiveness significantly. For a delay of 48 hours and a isolation efficacy of 50% (upper right corner) the app uptake must be around 75% to control the pandemic spread. Assuming a more realistic, yet still conservative, isolation efficacy of 70%, in combination of a four hour delay, around 40% app uptake is sufficient. The lower right panel shows that 90% effective isolation of cases with four hours delay should be sufficient to contain the pandemic, without any tracing of contacts. This result is in line with the model results shown in^3^, and indicates that at this moderate reproduction number the pandemic cannot be driven solely by infections from pre-symptomatic individuals. Figure S9 shows the same results as Figure S8, but for *R*0 = 2.7. We see that for this reproduction number it is not realistic to control the pandemic based on on digital contact tracing alone, since even for the highest isolation efficacy and lowest delay (lower left corner) the necessary uptake is around 90%.

**Figure S8:**
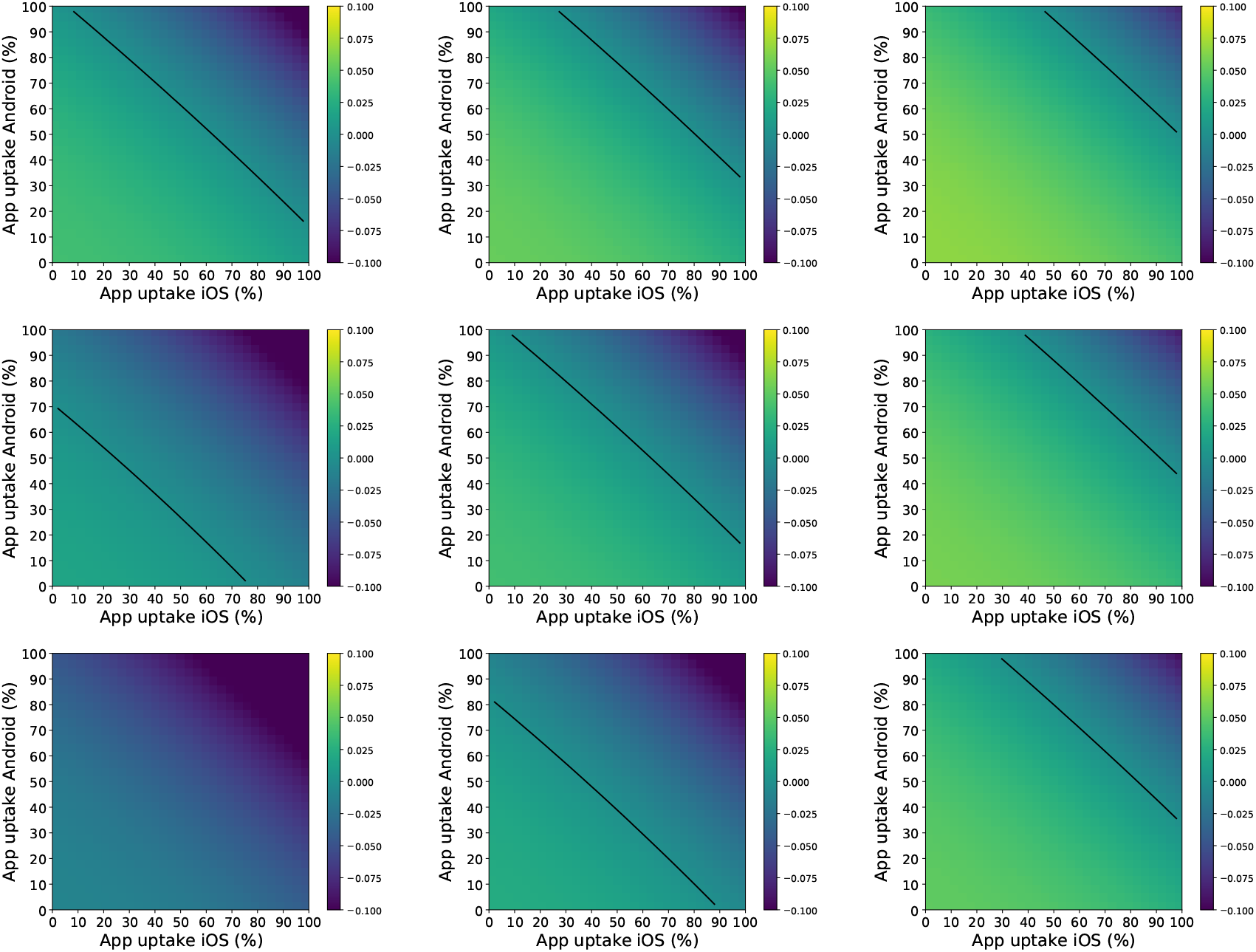
The figures show the exponential growth rate *r* as a function of app uptake, for different choices of model parameters. The reproduction number *R*0 = 1.5 for all plots. For the top row the efficacy of isolating infected cases is set to 50%, middle row 70%, and bottom row 90%. From left to right shows a delay of quarantining and isolation of 4, 24, and 48 hours, respectively.

**Figure S9:**
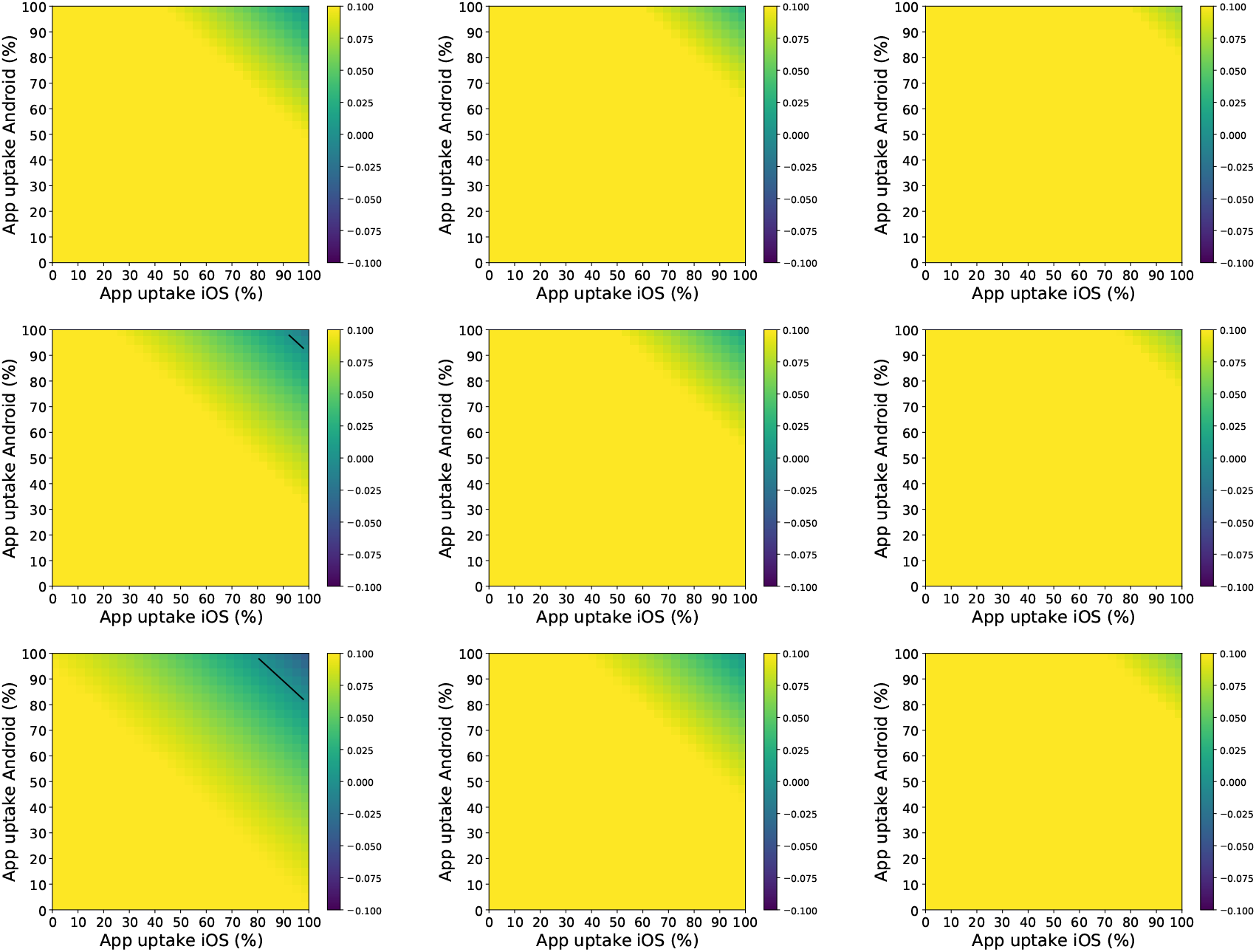
The figures show the exponential growth rate *r* as a function of app uptake, for different choices of model parameters. The reproduction number *R*0 is set to 2.7 for all plots. For the top row the efficacy of isolating infected cases is set to 50%, in the middle row 70%, and in the bottom row 90%. From left to right shows a delay of quarantining and isolation of 4, 24, and 48 hours, respectively.

## Acknowledgments

We are grateful to the Norwegian Institute of Public Health for allowing us to collect and use the anonymized Bluetooth contacts dataset. We would like also to thank Aslak Tveito for insightful discussions and suggestions and Maksim Kitsak for helpful discussions and comments.

## Data and Material availability

The data and scripts for the efficacy model, the impact of uptake on pandemic progression (i.e. Figures 2 and 4) and analysis of the ENS are available at^60^.Correspondence and requests for materials should be addressed to Ahmed Elmokashfi.

## Footnotes

1 Smittestopp had an age limit of 16.

